# Understanding bias when estimating life expectancy from age at death: A simulation approach applied to Morquio Syndrome A

**DOI:** 10.1101/2020.10.25.20219311

**Authors:** Xue Yin, Jaeil Ahn, Simina M. Boca

## Abstract

**Background:** Life expectancy can be estimated accurately from a cohort of individuals born in the same year and followed from birth to death. Due to the difficult and time-consuming nature of following a cohort prospectively, life expectancy is often assessed based on death data, which may lead to potentially biased estimates. This is more likely to be a problem in rare diseases such as Morquio syndrome A.

**Method:** To investigate how accurate the estimation of life expectancy is using death data, we simulate the survival of individuals with Morquio syndrome A under four different survival scenarios. In each scenario, we estimate the mean and median survival times within a defined period and compare them with the true life expectancy.

**Results:** When life expectancy is constant during the entire period, using death data does not result in a biased estimate of life expectancy. However, when life expectancy increases during the follow-up period, using only death data leads to a substantial underestimation of life expectancy.

**Conclusion:** Life expectancy can change over time, along with changes in the environment and/or biomedical innovation. When the life expectancy is increasing — as is often expected to be the case in rare diseases — estimating it based on contemporary death data will result in a downward bias. Therefore, it is crucial to understand how estimates of life expectancy are obtained and to interpret them in an appropriate context, and to assess estimation methods within a sensitivity analysis framework, similar to the simulations performed herein.

## 1 INTRODUCTION

Life expectancy is generally defined as the amount of time an individual can expect to live from birth; thus, it may refer to either the mean life expectancy or the median life expectancy. There are two main approaches to estimating life expectancy: cohort and period life expectancy. Cohort life expectancy is the average length of life from an actual cohort of individuals born in the same period. Since it is challenging to follow up individuals from birth to death prospectively, life expectancy is often evaluated using the average length of life in a hypothetical cohort of individuals who are assumed to have been born and died with the mortality rate observed in the same period[1, 2], known as period life expectancy.

The use of period life expectancy may result in a biased estimate, however, if the true life expectancy changes over time. In particular, it is conceivable that the life expectancy of individuals with certain diseases has been increasing recently due to improved diets, environmental changes, or biomedical innovation [3].

By definition, estimating life expectancy based on contemporary or past death records excludes living individuals, meaning that any recent changes in survival cannot be accounted for by methods like period life expectancy [4]. This problem is exacerbated in the case of rare genetic diseases such as Morquio syndrome A (also known as mucopolysaccharidosis type IVA or MPS IVA), which has an estimated birth prevalence (incidence) between 1 in 71,000 and 1 in 179,000 [5]. In the case of such a rare genetic disease, it will be more difficult to identify a sufficient number of individuals to follow from birth to death. In addition, estimates of the general population’s life expectancy will not be applicable due to the generally severe course of these disorders. Moreover, given the potentially impactful benefits of early diagnosis and improvements in general care[6], the life expectancy of individuals with rare diseases may be more likely to have increased in recent years compared to the life expectancy of the general population, leading to greater discrepancies when using period life expectancy estimation.

In the current study, we investigate how using data on only deceased individuals can lead to biases when estimating life expectancy, exemplified by MPS IVA. MPS IVA is a Mendelian autosomal recessive disease, with affected individuals having two abnormal copies of the N-acetylgalactosamine-6-sulfate sulfatase gene [5], one inherited from each parent [7]. Unlike non-Mendelian diseases, where life expectancy is often estimated from the time of diagnosis, for genetic diseases like MPS IVA it is usually considered from birth.

MPS IVA is caused by the deficiency of the enzyme N-acetyl-galactosamine-6-sulfatase, leading to the accumulation of glycosaminoglycans in body tissues, with multiple body systems being affected [8, 9]. Common symptoms found in affected individuals are short stature and skeletal dysplasia with bone deformity [10]. Their digestive, cardiovascular, and respiratory systems may also be affected, and they may have difficulties in hearing and speaking. The onset of disease symptoms is often between 1 and 3 years old [11].

Life expectancy for individuals with MPS IVA was previously estimated based on deaths that occurred over 36 years — between 1975 and 2010 — in the United Kingdom, using data collected by the Society for Mucopolysaccharide Disease [12]. A total of 27 deaths were recorded, the dataset including date of birth, gender, date of death, and primary cause of death. The mean age at death was 25.3 years (range of 3.08-75.32 years), with the most frequent primary cause of death being respiratory failure.

In the current study, we conduct simulations to assess different biases that can affect the estimation of life expectancy from mortality data, as was performed in [12]. As these estimates may be used by individuals with MPS IVA and their caregivers and providers to understand disease prognosis and plan possible interventions, it is crucial to provide a more nuanced interpretation and increase understanding of potential biases.

## 2 METHODS AND RESULTS

### 2.1 Reproducing the 2014 Analysis

We first attempted to reproduce the analysis in [12] by fitting a simple linear regression model. We used age at death (*y*) against the year of death (*x*), considering changes in the life expectancy of individuals with MPS IVA from 1975 to 2010. This analysis showed that affected individuals had a slow linear growth over time in life expectancy, *y* = − 1424.9 + 0.7368*x* (*R*^2^= 0.0963). Using the data provided in [12], we obtained a similar, but not identical result: *y* = −1344.7209 + 0.6862*x* (*R*^2^= 0.0922). While there is a slight discrepancy between the data presented in [12] and the data used by those authors to fit the linear model, we do not anticipate this to cause a major change in results.

### 2.2 Simulation scenarios

We simulate the birth and death of individuals with MPS IVA for four different scenarios, summarized in Figure 1 and described in more detail below.

**Figure 1:**
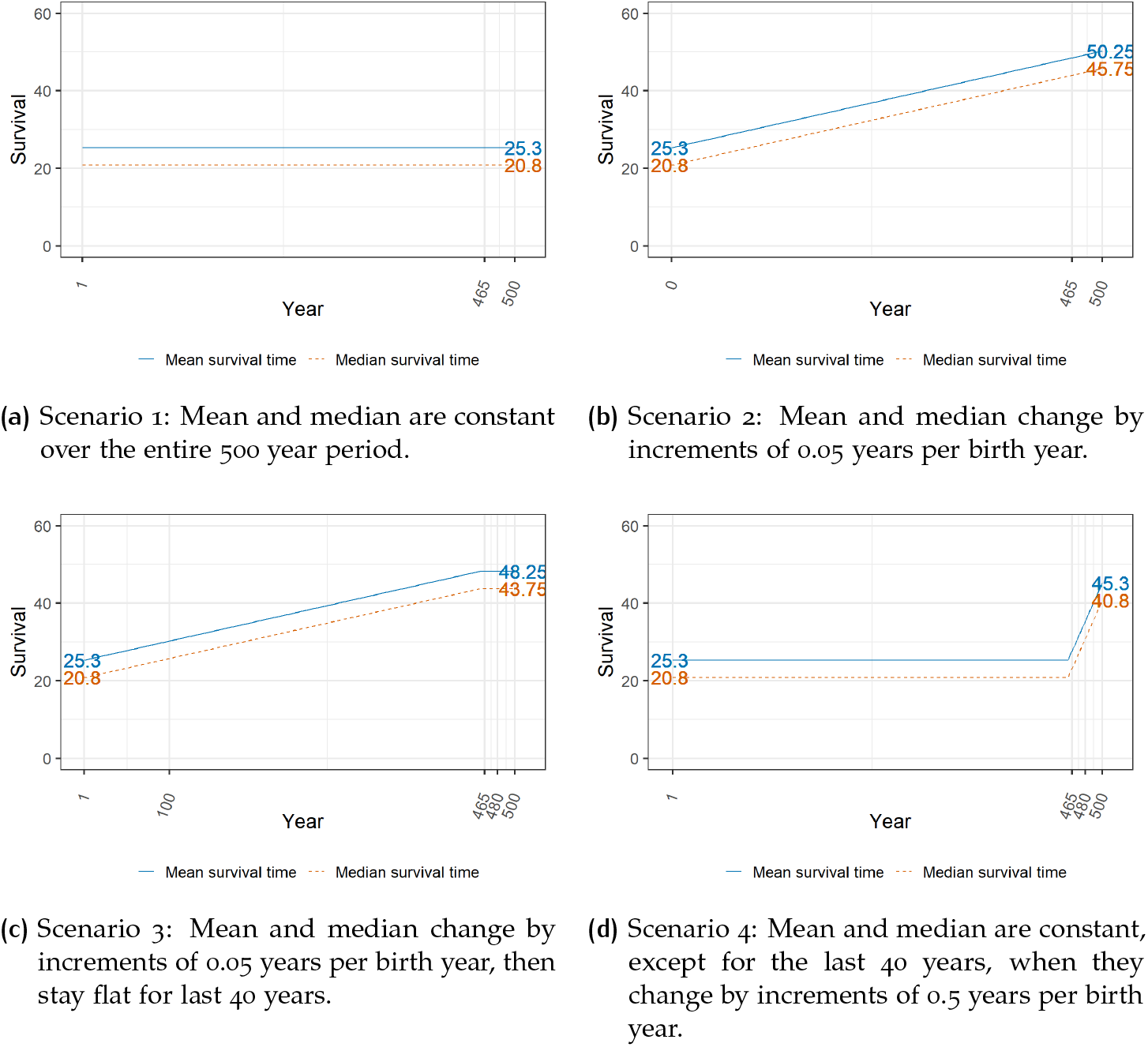
True mean and median values for the different scenarios. The solid blue line represents the true mean survival time. The orange dashed line represents the true median survival time.

We generate survival times based on the Weibull distribution, over 500 years, assuming a constant birth prevalence of one individual with MPV IVA born every year. We repeated 1,000 simulations to obtain summary statistics. In order to mimic the MPS IVA data collection in [12], we focus on the life expectancy for individuals born in the last 36 years — between years 465 and 500 — of the simulation. Thus, we discard the majority of the simulated individuals. We considered 500 years of forward simulation time to ensure that we would obtain a large enough number of deaths in the period [465, 500] to enable us to proceed with our analyses. In all scenarios, we consider that individuals born in year 1 have the same mean and median survival as in [12] (Mean=25.3, Median=20.8). We use the *simsurv* function from the R package *simsurv_0*.*2*.*3* to conduct all the simulations.

#### 2.2.1 Scenario 1: constant life expectancy

The first scenario, shown in Figure 1a, concerns a situation where the life expectancy is constant over 500 years. This implies that there is no change in treatment or environment that will influence the life expectancy of individuals with MPS IVA during the entire 500 year period. We generate survival from the Weibull distribution with scale parameter *λ* = 27.46 and shape parameter *k* = 1.32, which are obtained by using the mean of 25.3 years and median of 20.8 in the equations:

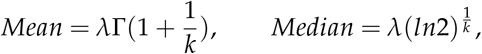

where Γ is the gamma function, and solving for *λ* and *k*. The density of this distribution is shown in Figure 2.

**Figure 2:**
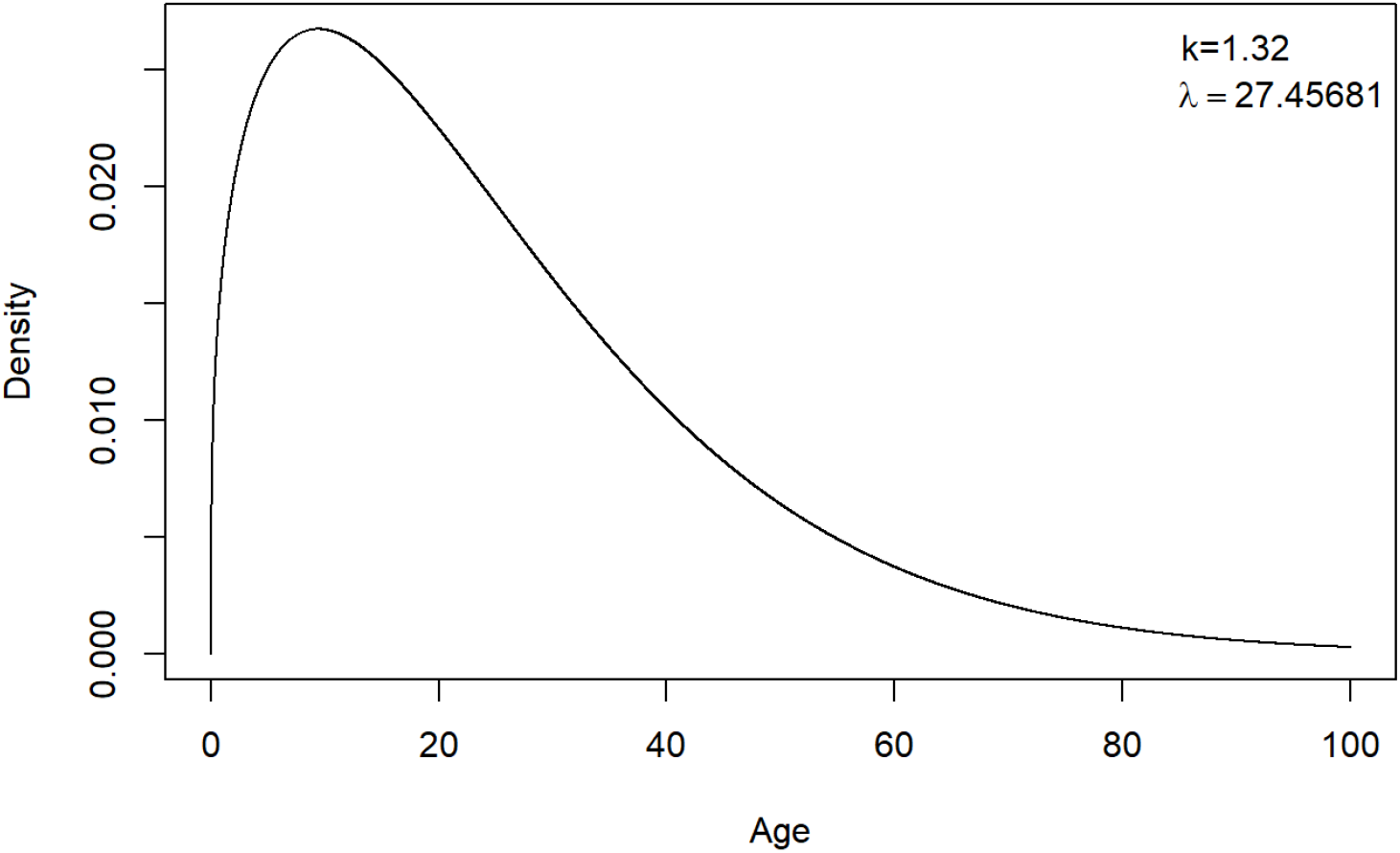
The density of the Weibull distribution used for generating survival times for the first simulation scenario, which considers constant mean and median survival of 25.3 and 20.8 years respectively, over a 500 year period.

#### 2.2.2 Scenario 2: Gradually increasing life expectancy

Contrant to the first simulation scenario with constant life expectancy, in Figure 1b, we assume that life expectancy increases linearly over time. This may reflect a situation where the treatment or care that individuals with MPS IVA receive has been consistently improving each year. This situation can occur when the standard of care gradually improves over time, without the introduction of an extremely effective intervention that dramatically changes the natural history of the disease. Here we set the mean and median survival to increase by 0.05 years each year, which means that for a simulated individual born in year 410, the mean and median survival time will be 50.25 and 45.75 years, respectively.

#### 2.2.3 Scenario 3: Gradually increasing life expectancy that later stabilizes

An alternative scenario is that life expectancy may improve up to a certain year, then stabilize without further improvements, for example if a treatment or a care protocol is refined up to a certain point, then stops improving, but continues being utilized and effective. For instance, while surgery complications appear to have reduced over time [12], there may be a point past which surgery can no longer further improve outcomes for individuals with MPS IVA. We model this in the third simulation scenario, in a situation where life expectancy increases for the first 460 years, then stabilizes in the last 40 years, as visualized in Figure 1c. The mean and median survival increases by 0.05 years per birth year until year 460, meaning that simulated individuals born in the year 460 have a mean and median survival of 48.25 and 43.75 years, respectively. Life expectancy is then constant until the year 500.

#### 2.2.4 Scenario 4: Constant, then increasing life expectancy

In the last scenario, we assume that life expectancy is stable for the first 460 years and only increases in the last 40 years, as shown in Figure 1d. This can be a situation where a new treatment appears that leads to a gradual improvement, perhaps by being introduced to increasingly younger individuals. We assume this treatment is more effective than the treatments from the second and third scenarios, increasing mean survival by 0.5 years per year starting in the year 460, meaning that individuals born in the year 500 have a mean survival time of 45.3 and a median survival time of 40.8 years.

### 2.3 Methods for estimating life expectancy

We consider a variety of approaches to estimate the life expectancy of individuals who were alive at some point within the last 36 years of the simulated time period. The period life expectancy approach is equivalent to [12], using only data on the individuals who died during that period to estimate mean and median life expectancy. For the cohort approach, we use the full survival times of individuals born between years 465 and 500; however, in practice, these would not be available if the analysis was performed at year 500.

We also consider the Kaplan-Meier (KM) method to estimate the median survival in the presence of censored data[13], which allows us to include partial survival times as censored times for individuals who are still alive at year 500. We estimate median life expectancy for both retrospective (R) sampling, where we consider individuals who died since year 465 and prospective (P) sampling, where we instead follow up individuals who were born since year 465. Estimating the median survival time of all individuals who died between years 465 and 500 without censoring at year 500 via the KM approach is equivalent to the period life expectancy approach. If censoring is considered at year 500, the resulting survival times are often heavily censored, so we also consider KM estimation that weights censored individuals, which reduces their influence. In particular, we considered both weighting censored individuals by 0.1 — due to this factor working well in practice — and by the percentage of uncensored individuals (uncensored percentage) among all sampled individuals[14]. For prospective sampling, the KM estimation censors all individuals still alive at year 500.

### 2.4 Simulation results

Results from the 4 simulation scenarios — giving the true mean and median life expectancies and the estimates from the period approach — are presented in Table 1 and Figure 3. Table 1 presents the summary results, along with the mean period and cohort estimates, approach, while Figure 3 also shows the individual results for each of the 1,000 simulations runs for the period approach. For scenarios 2-4, where the life expectancy was not constant over the entire period, the “true values” are given by the average mean and median values over the last 36 years.

**Table 1:** True mean and median survival and average estimated mean and median survival across 1000 simulation runs, for each of the 4 simulation scenarios, using both the period and cohort estimation approaches. The true values represent averages over the last 36 years for each scenario. The period and cohort approaches average the corresponding estimates over 1000 simulation runs. The numbers in parentheses represent the standard deviations across the 1000 simulation runs.

| Scenario | Mean survival time |  |  | Median survival time |  |  |
| --- | --- | --- | --- | --- | --- | --- |
|  | True value | Estimates |  | True value | Estimates |  |
|  |  | Period | Cohort |  | Period | Cohort |
| 1 | 25.30 | 25.24 (2.92) | 25.35 (3.28) | 20.80 | 20.75 (3.68) | 20.89 (3.68) |
| 2 | 49.37 | 45.91 (4.18) | 49.52 (4.95) | 44.88 | 42.10 (6.02) | 44.96 (6.02) |
| 3 | 48.25 | 45.67 (4.2) | 48.37 (4.89) | 43.75 | 41.83 (5.91) | 43.77 (5.91) |
| 4 | 36.55 | 27.93 (3.16) | 36.63 (4.10) | 32.05 | 23.90 (4.99) | 31.96 (4.99) |

**Figure 3:**
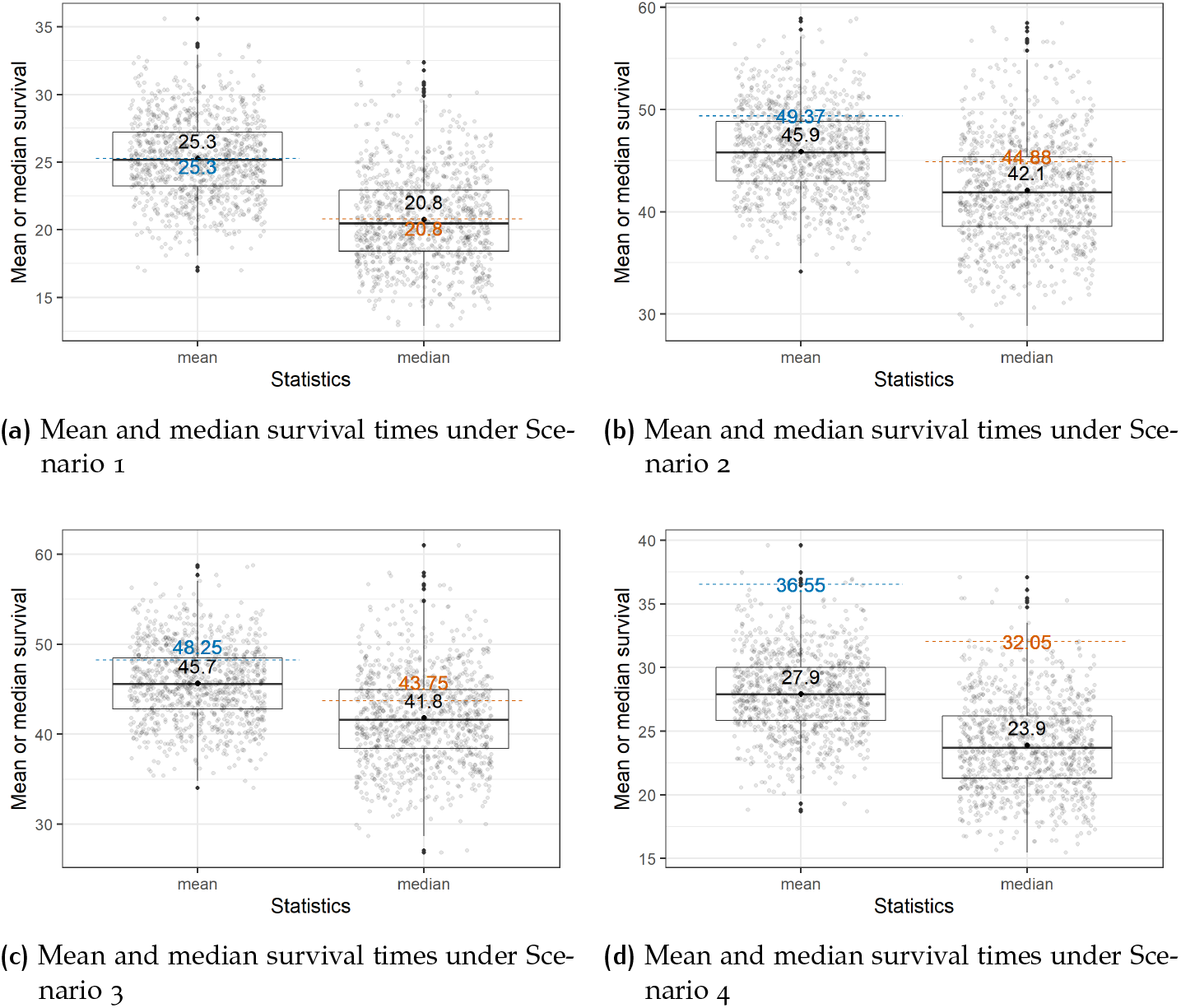
Boxplots of estimated period mean and median survival times for simulation scenarios 1-4. The blue dashed line is the average of the true mean survival time for the last 36 years. The orange dashed line is the average of the true median survival time for the last 36 years. Each grey point represents the result of a single simulation run, with the means and medians estimated via the period approach, using only the simulated individuals who died in the last 36 years within that run. The black point is the average value of the grey points over 1000 simulation runs.

For the first scenario of constant life expectancy, the estimated period means and medians from the 1,000 simulation runs are close to the true values, with no notable bias. However, with the departure from the constant life expectancy assumption in the remaining three scenarios, the estimated period mean and median estimates consistently show downward biases. The estimated cohort mean and median survival estimates — which use all individuals born since the year 465 — are close to the true values in all four scenarios.

The average KM estimates for the four simulation scenarios are shown in Table 2. The estimate using only individuals who died between years 465 and 500 (with no censoring at year 500) is the same as the period life expectancy estimate, but we include it here as well for comparison with the other KM estimates. Considering the individuals who are still alive at year 500 as censored, the KM approach overestimates the median life expectancy under all scenarios. The KM approach weighting censored individuals by 0.1 — which reduces the influence of heavy censoring — yields median survivals closer to the true medians than the other KM methods. When weighting by the uncensored percentage among those individuals, the KM estimator performs worse than when weighting by 0.1 but is closer to the true median compared to the versions that do not weigh the censored data. For the prospective approach, which considers individuals born since year 465 and censored at year 500, the KM method cannot be fit in Scenarios 2 and 3, because on average, only around 5 individuals die within the period [465, 500] years, the remainder being censored. As an additional sensitivity analysis, we also considered individuals born since year 500. In that situation, there were approximately 27 out of 51 uncensored individuals in Scenario 1 on average, 13 in Scenarios 2 and 3, and 22 in Scenario 4; the KM method successfully estimated median survival, which slightly differed from the average true median for the last 50 years.

**Table 2:**
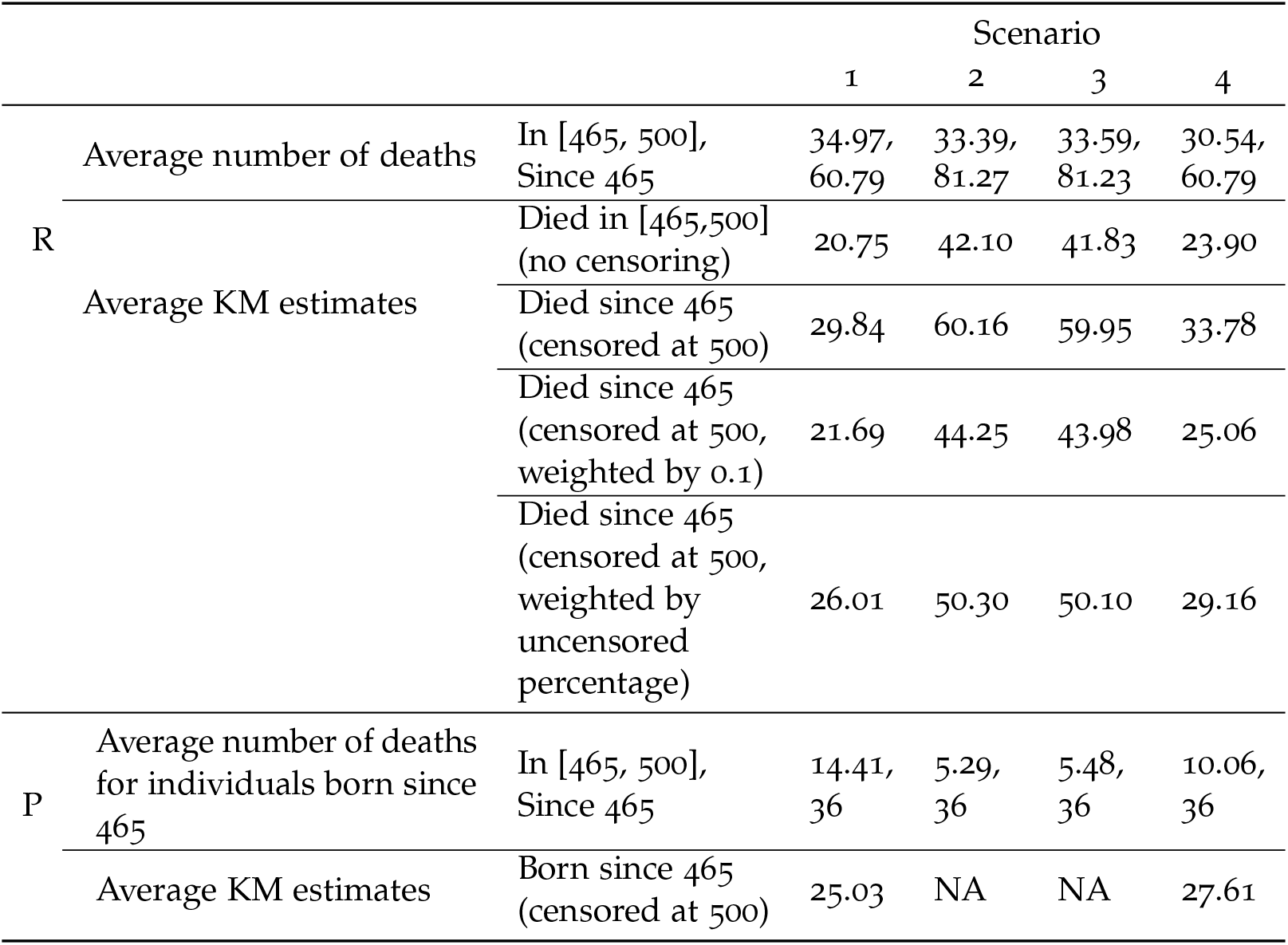
Average number of deaths in the time period [465, 500] years and Kaplan-Meier estimates of the median survival time over the four simulation scenarios, over 1,000 simulation runs in each scenario. R = retrospective sampling (considering individuals who died since a certain year), P = prospective sampling (considering individuals who were born after a certain year). NA = estimation not possible due to small number of events.

### 2.5 Reproducibility

All the analyses in this manuscript are reproducible, with the code and data available at github.com/xueyin97/Bias-estimating-life-expectancy-Morquio.

## 3 CONCLUSIONS AND DISCUSSION

We investigated the problem of estimating life expectancy from data on age at death, focusing on Morquio syndrome A, using data provided in [12]. We initially replicated the primary analysis from [12], obtaining similar, though not identical results. We proceeded with simulating data based on summary statistics of the provided data, assuming known trends of survival over time.

Simulation studies under four conceivable scenarios were conducted to better understand the magnitude and types of biases that occur when only data on deaths are considered in estimating life expectancy. Firstly, we simulated the scenario where life expectancy is constant for 500 years. We found that the estimated means and medians that only use the death data — in other words, the period approach — performed well in this case, with no apparent bias. It is known that, under constant mortality, mean period life expectancy is the same as average age at death [4].

The scenario assuming constant survival over time fails to reflect the advances in medical practices and technology. Changes in general care and treatment of MPS IVA are expected to lead to a potential rise in life expectancy, which may have already started but was not captured in [12]. Indeed, improved treatments such as enzyme replacement therapy and hematopoietic stem cell therapy have recently started being used for MPS IVA and may eventually have a positive impact on survival [15]. Even in the absence of specific treatments, standards of care have improved along with advancements in medical devices and techniques, leading to a potential increase in life expectancy [16]. For example, [12] found that although surgery complications had previously been one of the major causes of death for MPS IVA, since 1990, they only resulted in a small percentage of deaths, which might be a consequence of improvements in surgical and post-surgical care.

To address the likely scenarios of life expectancy improving over time due to the changes in standard of care or the improvement in medical treatment, we considered three additional scenarios, representing a mix of stable and increasing life expectancy, which we considered to be potentially more realistic for the case of a rare disease like MPS IVA. In all the non-constant life expectancy scenarios, period life expectancy substantially underestimates both the mean and the median life expectancy. Thus, unless the life expectancy is relatively stable over time, the approach used in [12] will not accurately assess life expectancy.

In contrast with period life expectancy, which assumes stable mortality rates over the period of interest, cohort life expectancy allows for possibly time-varying mortality rates. Under the first scenario, for which the life expectancy is stable as the mortality rate is constant over time, the period life expectancy, therefore, equals the cohort life expectancy[4]. In the remaining scenarios, for which life expectancy is increasing as mortality rates decrease, the period approach underestimates the true life expectancy, while the cohort approach leads to unbiased estimates. However, the cohort approach is generally impractical, especially for rare diseases like MPS IVA, which make it more difficult to find and follow up a large enough number of individuals. Moreover, with a cohort approach, there is always a lag between the life expectancy estimated on a cohort for which everyone is deceased and the life expectancy for an individual born at the present time. We also considered the KM method — which allows for the inclusion of individuals who are still alive as “censored data” — to estimate the median survival. Using KM estimation for the median survival did not eliminate bias, although it changed its direction. The KM estimate for individuals who died between the years 465 and 500, censoring individuals still alive at year 500, overestimated true survival in all four scenarios. This may be expected given the high rate of censoring — on average, 58% of individuals were censored under the first scenario, 41% percent under the second and the third, and 50% for the fourth scenario — and the tendency of the KM estimator to be anti-conservative in these conditions [17, 18]. To overcome this problem, we considered using weights for the censored data. Weighting the KM median by 0.1 leads to greatly improved estimates, but since it was chosen based on the known “ground truth” in simulated examples, this cannot be recommended in general. Approaches also exist to weight the censored data by the percentage of the uncensored individuals among all sampled individuals [14, 18, 19]. This still leads to an overestimation of the median survival, but provides a better option than the unweighted KM method. We also considered a prospective sampling design, following up individuals who were born since year 465, but censoring them at year 500. In this case, since we only have 36 individuals born since 465 due to our assumptions, it becomes impossible to obtain an estimate in two of the scenarios, as the average number of deaths for those individuals in the period [465, 500] years is below 6.

Our study has a number of limitations, the most important being that the simulated datasets are based on simplified assumptions. There is not enough knowledge of changes in the natural history of Morquio syndrome over time to build detailed models, leading to our choice of a number of scenarios as a de facto sensitivity analysis for comparing the performance of various approaches to estimate life expectancy. While in general, life expectancy is expected to increase over time, we note that the current COVID-19 pandemic situation may lead to a reduced lifespan for individuals with underlying respiratory conditions [20], including MPS IVA. To estimate the life expectancy more accurately, we need to consider contemporary changes in the standard of care and medical treatment and the possibility of unforeseeable and unfavorable events, which may include pandemics.

When estimating life expectancy, especially in rare diseases, using the cohort approach with long-term data will be more accurate than using the period approach, but this design also has drawbacks and results in lagged estimates. If using a period approach based on death data, we must assume that the result will not reflect the current life expectancy and will probably underestimate it in the case of improvements in treatment or general care. An alternative is to also include data on individuals who are still alive by employing a weighted KM method; while this approach appears to lead to anti-conservative biases, it performs better than the unweighted KM method. Future methods will require a balance of these aspects, potentially by incorporating certain reasonable assumptions on the underlying life expectancy and considering extensive sensitivity analyses.

## Data Availability

The dataset we used had previously been published in:
C. Lavery and C. Hendriksz. Mortality in Patients with Morquio Syndrome A. JIMD Reports, 15, 2014.

https://link.springer.com/chapter/10.1007/8904_2014_298

https://github.com/xueyin97/Bias-estimating-life-expectancy-Morquio

## REFERENCES

[1] J.R. Goldstein and K.W. Wachter. Relationships between period and cohort life expectancy: Gaps and lags. Population Studies, 60:257—269, 05 2006.

[2] H. Shryock, J. Siegel, and E Larmon. The methods and materials of demography. U.S. Bureau of the Census, 2 edition, 1973.

[3] F.R. Lichtenberg. The impact of biomedical innovation on longevity and health. Nordic Journal of Health Economics, 5:45—57, 2017.

[4] M. Guillot. Period versus cohort life expectancy. In Rogers R. and Crimmins E., editors, International Handbook of Adult Mortality, pages 533–549. Springer Netherlands, 2011.

[5] R.M. Leadley, S. Lang, K. Misso, and et al. A systematic review of the prevalence of Morquio A syndrome: challenges for study reporting in rare diseases. Orphanet Journal of Rare Diseases, 9, 2014.

[6] K. Bhattacharya, S. Balasubramaniam, Y. Choy, and et al. Overcoming the barriers to diagnosis of morquio a syndrome. Orphanet Journal of Rare Diseases, 9, 2014.

[7] A. Morrone, A. Caciotti, R. Atwood, and et al. Morquio a syndrome-associated mutations: A review of alterations in the galns gene and a new locus-specific database. Human mutation, 35, 09 2014.

[8] J. Singh, N. Ditterrante, P. Niebes, and Tavella D. N-acetylgalactosamine-6-sulfatase in man: abasence of the enzyme in Morquio disease. J Clin Invest, 57:1036—1040, 1976.

[9] J. Glössl and H. Kresse. Impaired degradation of keratan sulphate by Morquio A fibroblasts. The Biochemical journal, 203,1:335–338, 1982.

[10] J. Wraith. The mucopolysaccharidoses: a clinical review and guide to management. Arch Dis Child, 72(3):263—267, 1995.

[11] A. Montaño, S. Tomatsu, G. Gottesman, M. Smith, and T. Orii. International Morquio A Registry: clinical manifestation and natural course of Morquio A disease. J Inherit Metab Dis, 30(2):165—174, 2007.

[12] C. Lavery and C. Hendriksz. Mortality in Patients with Morquio Syndrome A. JIMD Reports, 15, 2014.

[13] E. Kaplan and P. Meier. Nonparametric estimation from incomplete observations. Journal of the American Statistical Association, 53(282):457–., 1982.

[14] M. Shafiq, S. Shah, and M. Alamgir. Modified weighted kaplan-meier estimator. Pakistan Journal of Statistics and Operation Research, 3(1)):39–44, 2007.

[15] S. Tomatsu, K. Sawamoto, T. Shimada, and et al. Enzyme replacement therapy for treating mucopolysaccharidosis type IVA (Morquio A syndrome): effect and limitations. Expert Opin Orphan Drugs, 3, 2015.

[16] J.D. Tobias. Anesthetic care for the child with Morquio syndrome: general versus regional anesthesia. Journal of Clinical Anesthesia, 11:242—246, 1999.

[17] M. Noordzij, K. Leffondré, K. Stralen, and et al. When do we need competing risks methods for survival analysis in nephrology? Nephrology Dialysis Transplantation, 28:2670–2677, 11 2013.

[18] K. Shoaib, A. Hamdi, and A.L. Ahmed. A compression of Kaplan Meier vs. weighted Kaplan-Meier in comparing estimation of heavy censoring data. American Scientific Research Journal for Engineering, Technology, and Sciences, 36:211–223, 2017.

[19] A. Zare, M. Mahmoodi, K. Mohammad, and et al. A comparison between Kaplan-Meier and weighted Kaplan-Meier methods of five-year survival estimation of patients with gastric cancer. Acta Medica Iranica, 52(10):764–7, 2014.

[20] A. Clark, M. Jit, C. Warren-Gash, and et al. Global, regional, and national estimates of the population at increased risk of severe COVID-19 due to underlying health conditions in 2020: a modelling study. The Lancet Global Health, 8(8):e1003.–e1017, 2020.

